# Creating a novel digital intervention to bridge the organizational and personal health literacy gap by teaching the way patients learn: a process and application study

**DOI:** 10.1101/2025.02.16.25322112

**Authors:** Maud Joachim-Célestin, Nishita Matangi, Lizbeth Rivas, Nikhil R. Thiruvengadam, Susanne B. Montgomery

**Author notes:** **Corresponding author:** Maud Joachim-Célestin, MD, Dr.PH, CHES P. O. Box 20, Loma Linda, CA 92354 **E-mail:**.

## Abstract

Low literacy is on the rise in the USA and is one of the independent predictors of poor health outcomes. While much written health information is now adapted to lower reading levels, more needs to be done to address the gap between material content and target consumers’ comprehension ability which often is more complex than simple reading level adjustments. This paper describes the process of developing a culturally, context and literacy-aligned intervention that promotes and supports preventive lifestyle behaviors among low-income Latino women (Latinas).

Focus group discussions and key-informant interviews were conducted to identify needs, barriers and beliefs of low-income Latinas regarding obesity, healthy eating and physical activity. A simple literacy-aligned, culturally-appropriate and socio-economically acceptable (LACASA) framework and intervention were then created through an academic-community partnership: medical professionals and students, community health workers and researchers.

The curriculum – which included minimal and easy-to-read written material and power point presentations - was piloted and revised before being launched. The resulting lifestyle curriculum for high-risk low-income Latinas was well-received and was a good fit for the priority population.

Creating a program using the LACASA approach requires an interdisciplinary team to invest time working with key members of the priority population and a commitment to adjusting materials to the group’s literacy level and its cultural and economic realities. Programs created to serve low-income individuals with limited literacy cannot rely on pre-created curricula. Instead, these must be re-evaluated and adjusted to address both content core principles and specific contexts of the priority population.

---

In the USA, the number of individuals with limited literacy is on the rise with more than 30 million people unable to read or write above third grade level and more than half of the population reading below the 6^th^ grade level. Even among those with a high school diploma, 1 out of 4 individuals is described as having limited functional literacy -which includes numeracy, reading and writing skills (U.S. Department of Health, 2020; Volaco A, et al., 2018). A recent assessment found that 21% of adults have low literacy levels, a phenomenon costing the country more than 2 trillion dollars a year (National Literacy Institute, n.d.). For a number of reasons, low-income populations tend to have one the highest rates of limited literacy with women and persons from ethnic and racial minorities being more affected: nearly half of African Americans and Hispanics have limited literacy and are often labeled “functionally illiterate” (Lueken et al, 2022; Price et al., 2018).

Limited functional literacy is a multifaceted problem, not only strongly associated with poverty (e.g., the need to leave school at a young age to provide income for the family, having to focus on surviving rather than education, violence in the community) but also a product of past and present US government policies that have led to educational inequity (including anti-literacy laws, distribution of resources in school districts, segregation laws) (American Anti-Slavery Committee, n.d.; Literacy Pittsburgh, n.d; Menendian et al., 2021). Moreover, a strong inverse correlation between literacy or reading/writing ability and a person’s health has also been described (Martin et al., 2011).

Indeed, functional literacy is considered one aspect of personal health literacy (Nutbeam, 1998), defined as the set of skills one needs to navigate health situations and make sense of the health information and services one is provided with (Centers for Disease Control and Prevention [CDC], 2023). Limited functional and health literacy have been shown to contribute to worst outcomes in a number of chronic conditions (diabetes, heart disease, mental health disease) – all of which require numeracy and literacy skills as well as understanding of complex concepts for self-management – resulting in higher mortality among groups most highly affected (Sanders et al., 2014). At the same time, for patients suffering from chronic diseases, literacy and educational attainment have been identified as important predictors of success in managing their disease, especially when individuals enroll in interventions (Bos-Touwen, . Jansen etal., 2018; van der Gaag, et al., 2023).

To communicate more effectively with patients, the American Medical Association and National Institute of Health created guidelines for written material targeting patients (US Department of Health and Human Services, 2021; Weiss, 2007). The Association of physicians for the underserved has even written material for individuals with “very low literacy” but even some of these materials have been found to still be challenging (Association of Clinicians for the Underserved, 2023). Still, written material shared with patients suffering from chronic diseases continues to be at a higher reading level than the recommended 6^th^ grade reading level (Han & Carayannopoulos, 2020). Even for patients with the most common chronic health conditions, available health education material is beyond the average reading level (Boutemen & Miller, 2023; Gajjar et al, 2024; Nash et al, 2023). While the Center for Disease Control and Prevention National Diabetes Prevention Program (NDPP) material is written for 6^th^ grade for English speakers and 5^th^ grade for Spanish speakers (Centers for Disease Control and Prevention, 2022), the program also encourages reading food labels, weight tracking, reporting (often using grids and graphs) - all skills that also require numeracy skills. Most self-management and preventive interventions continue to rely heavily on written material and self-monitoring skills (Painter et al, 2017) and there are currently few “low literacy” intervention programs available (Pignone et al, 2005; Sheridan, S. L. & et al., 2011; van der Gaag, et al., 2023). The latest standards of care for diabetes suggest adapting the intervention to “technology access, education, literacy, and numeracy skills” (ElSayed et al, 2023) but no specific instructions are given on how to successfully adapt the intervention to the literacy level of program enrollees.

With the COVID-19 pandemic, more programs are available online but, once more, the reading level, surpasses the comfort level of most Americans (Lee et al., 2022; Singh et al., 2022). Furthermore, access to and comfort with technology is often non-existent or minimal among some of the underserved populations (Lipari et al., 2019; Novin et al., 2019).

One of the Healthy People 2030 goals is to “increase the health literacy of the population” (2023) with an emphasis on health-related organizations educating providers, disseminating information at a reading level that is understandable for patients, and increase the proportion of patients’ personal health literacy to 4^th^ grade level. Thus, both organizational and individual health literacy are promoted (HHS, 2021d)

Besides being a barrier to understanding instructions, limited functional literacy is often a source of embarrassment, leading participants to feel ashamed, intimidated or ostracized during an intervention or a medical encounter (Easton et al., 2013). In one study patients reported never telling anyone they struggled with literacy, not even to their own family member. This type of shame can have an impact on patient relationships with providers and on patient decisions for care (Parikh et al., 1996; Lyons & Dolezal, 2024). Thus, individuals with low literacy are often unlikely to ask questions. Lastly, some may find it difficult to identify with those who deliver the intervention because their realities (living conditions, circumstances) are so misaligned (Sasson et al., 2015; Shelton et al., 2011).

Furthermore, different cultural contexts can add to an already complicated conversation leading to less understanding and, at times, more intimidation (Burki, 2020). These can be in the form of misinterpretation of body language or of certain behaviors, assumptions or comments to/about patients that are not culturally sensitive, resulting in less-than-ideal provider-patient communication and healthcare outcomes (Stubbe, 2020; Vandecasteele et al., 2024). With all these barriers, it is easy to understand why it may be difficult for someone with low literacy, especially of a different culture, to enroll in, and complete a needed intervention.

The person facilitating the instruction is therefore important. There is evidence that implementing a culturally appropriate intervention facilitated by community health workers (CHWs) can help bridge cultural and language barriers among many low-income persons. CHWs are often the most trusted healthcare representatives since they come from, and understand, the community patients live in. They are also well aware of resources available to address social determinants of health and barriers that community members face (Grant et al., 2017; Jacob et al., 2019). The most effective interventions are culturally appropriate (Stormacq, et al., 2019).

Employing community health workers to facilitate the process is critical as they provide a safe environment and reduce the potential intimidation that is likely during the learning process. As such, they can play a critical role in translational research and are well fitted to convey key messages to the community in an environment of trust.

A recent meta-analysis concluded that a combination of culturally competent healthcare strategies at both organizational and individual provider-patient level would be ideal. In that model, the individual level includes culturally and linguistically adapted materials, incorporation of culturally-specific concepts, linguistic and culturally matching staff, involvement of families and continuity of care and integrating community health workers in patient care. (Handtke,

Schilgen, & Mösko, 2019). Other models and strategies have also been described, including the logic model used for diabetes patients (CDC, 2018) to address the issue of interventions in this population, with some interventions having been created and implemented successfully among individuals with limited literacy

Individuals with limited functional literacy and low income tend to be already burdened and stressed, making the learning process challenging (Bekteshi, 2024; Ridley, 2020).

Therefore, requiring them to increase their literacy adds an additional burden (including that of acculturation) to individuals often already too burdened to learn due to multiple competing priorities. Instead, organizations offering services and information to these individuals must be willing to adapt to their needs, Focusing on more effective means of communication and facilitators that are a better fit may be especially critical, because of it may be unrealistic to provide culturally concordant staff as suggested by Handtke, Schilgen, & Mösko (2019).

However, material must be user-friendly for CHWs who themselves may require properly adapted didactic material that fits theirs as well as participants’ literacy levels. While ideal agents to deliver the program, it is too much to expect them to adapt the provided material “on the fly”. Furthermore, while recommendations have been made, no specific patterns have been identified to create material for use by CHWs without requiring program planners to start “from the ground up” every time. There is a need for a set of guidelines to inform the production of material that is literacy-aligned, culturally-appropriate and socio-economically acceptable (LACASA) for use by CHWs who lead self-management programs among individuals of similar cultural background with limited literacy and financial flexibility Clearly, carefully prepared and relevant materials should be available for participants and CHWs equally (Moffett et al., 2018).

In this article, we propose and describe the literacy-aligned, culturally-appropriate and socio-economically acceptable (LACASA) model After explaining the steps to creating and implement these instructional materials for a lifestyle intervention, we also describe a practical application of the process as implemented by Hispanic/Latino CHWs leading an intervention among low-income low literacy Latinas living in California (for purposes of this article we use the term Latino/a/x recognizing that many individuals prefer the term over the term Hispanic). Our hope is that the framework/constructs and applications shared will serve as a resource to increase organizational health literacy, thus reducing the health disparities gap that exists in the healthcare of those with low-literacy, limited financial flexibility and unique cultural needs.

## Methods

The steps to create a LACASA intervention broadly parallel the main phases of the (social marketing assessment and response tool) model, which applies the major principles of the Equity-Centered Health Communication Framework (CommunicateHealth, Inc., 2023), a framework recently developed as an expansion of the CDC “pink book”. Table 1 and Figure 1 provide an overview of the recommended process (steps and time frames). Most steps involved open communication with CHWs from the same cultural and socio-economic background as the population of interest. A community-based participatory approach has been shown to be crucial for culturally-appropriate materials, especially among individuals with limited literacy (Kreuter, 2003).

**Figure 1.**
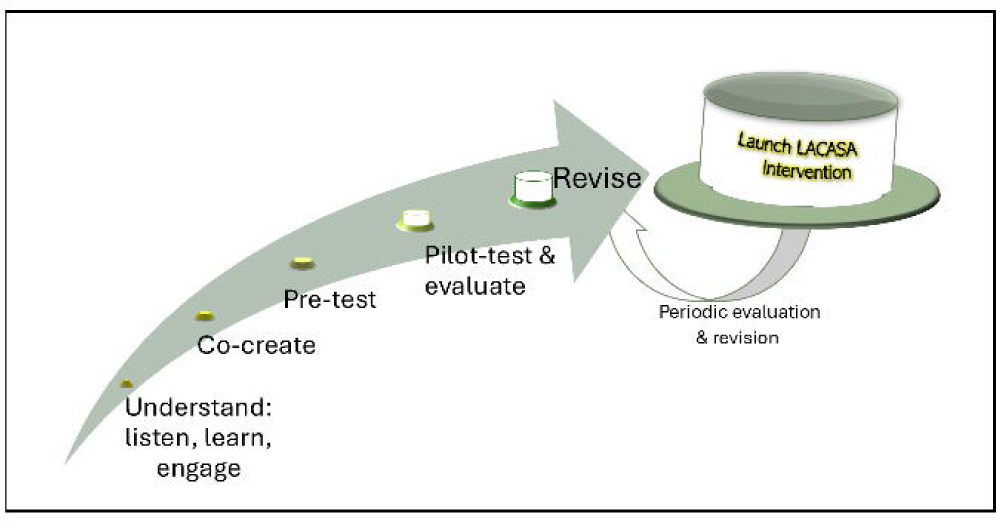
Graph depicting the steps used to create a literacy­ aligned, culturally-appropriate, socio-economically acceptable (LACASA) intervention

### Step 1: Understand – Listen, Learn, Engage

#### Tuning into the Local Culture and Finding Local CHW Partners

This step involves identifying beliefs and norms of the select population on the topic/condition to be addressed. To allow for unbiased ideas and avoid preconceived notions, this is done prior to literature review and by engaging in conversations with various community members in their “natural settings”. This intentional “immersion” in the community may be best done in concert with local CHWs. Field notes should be kept, and special focus should be placed on social etiquette (acceptable/unacceptable behaviors, currently used terms and communication styles).

#### Conducting Literature Search

A literature search is then conducted to confirm or add to the understanding of the information gained to date. Based on gathered information, a list of questions is then created to engage in the subsequent qualitative work (key informant interviews and focus groups) which will clarify what information is applicable to this specific community.

#### Facilitating Focus Group Discussions and Interviews

The goal of this step is to gain a better perspective on cultural and other relevant realities by hearing directly from members of the population of focus. It involves conducting and recording focus group discussions (FGDs) – ideally no more than ten people at a time - with community health workers and possibly other key members of the community. Conducting “key informant” interviews may help detect issues that people may only be comfortable sharing privately. It’s also best to conduct these in trusted and easily accessible facilities for the population of interest. Each participant is given a small compensation as a token of appreciation for their time.

#### Summarizing Target Population Norms, Beliefs, Wants and Needs

Recordings of focus group discussions and interviews are then transcribed, coded and analyzed for emerging themes. Information gathered from these, from the field notes and from the literature review is then synthesized with a special emphasis on identifying norms, beliefs and expressed wants and needs of the population in regards to the specific topic/condition.

### Step 2: Co-create Key Intervention Components

Once there is a good understanding of views and challenges of the population of interest, an interdisciplinary team is carefully selected to create intervention components.

#### Gathering an Interdisciplinary Team

To provide a comprehensive and relevant program application, the ideal team must include a good representation from the selected population including community health workers or equivalent and someone with personal experience with addressing the topic/condition. The goal is for the team to look at the summary of all the gathered information and organize it to determine 1. Key material content; 2. Design, presentation, and type of didactic style/materials+ to be used; 3. Best setting/times to hold the intervention.

#### Selecting an Intervention Framework

Based on the information gathered about the population of interest, the team then proceeds to select a short list of key intervention components to be emphasized, and then to identify an existing intervention that incorporates most, if not all, of these key concepts. It could be an evidence-based intervention that is adaptable to a low income, low literacy population and requires minimal numeracy ability.

#### Developing Content of New Intervention

From the very beginning, discussions (and later interaction with program participants) must be in keeping with “popular education” and “adult learning” didactic methods, approaches which have been shown to work well with low-literacy low-income populations. The popular education approach is based on concepts popularized by Paul Freire whereby no one is “the expert”, but individuals involved in the process are co-learners equally engaged in the discussion. A facilitator only helps fellow team members come up with their own conclusions (Toro-Morn, 2012). Adult learning, popularized by Malcolm Knowles, assumes that participants are highly motivated, able to choose their own learning style, learn and contribute best using their past and current experiences. They only need some encouragement and positive feedback (Knowles, 1968; Wiggins, 2011). The more interactive the process, the better.

It is important that, as much as possible, key messages of the selected intervention framework are adapted or expanded so as to be consistent with and reinforce (vs reject) participants’ cultural norms/traditions. Tangible solutions to participants’ perceived needs/barriers requiring minimal decision-making efforts (from participants) will help facilitate informed decision-making (Kennedy et al., 2017; López et al., 2016). Lastly, small surprises should be planned to be given out at each session to keep participants engaged.

#### Developing New Curriculum Material

After developing key intervention content, team members set out to create didactic material and participants’ material, the latter being ideally a workbook that comes alive for members of the select population. A way to systematically ensure reading comprehension and relatability at all reading levels is to use material that requires minimal literacy and numeracy and includes illustrations/pictures that participants can relate with (CommunicateHealth, Inc., 2023). Recommended guidelines for material written for individuals with limited literacy - minimal, short and simple words and sentences, bullet lists, universal icons, many pictures, colorful presentations and visual aids - should be followed (Baig, et al., 2012; New York University, 2024; Wolff, 2009). Letters should also be large and clear enough to be easily deciphered without reading glasses (several may have poor eyesight or inadequate reading glasses, if any). Color selection and color combinations must also be carefully selected as these have meanings and convey subliminal messages that vary from culture to culture (Racoma, 2019; De Bortoli & Maroto, 2001; Interactive Design Foundation, 2021; Color Meanings, n.d.). Whenever photographs of individuals are used, these should be representative of members of the population of interest (age, physical appearance, clothing, activities, settings).

To be culturally appropriate, the material must be visually appealing to the target audience and relatable. Individuals must relate and want to engage with it (Kreuter, 2003).

### Step 3: Pre-test and Fine Tune Material

#### Pre-testing Material for Clarity, Relevance and Literacy/Cultural Appropriateness

At the end of the brainstorming and material development phase, it is important that CHWs from the same cultural background facilitate a “teach-back” session using newly created didactic material (e.g., flipchart, facilitator guide) and participant workbook. Meanwhile, fellow team members assess/grade the process, simplicity, clarity, feasibility, and cultural relevance of the session facilitated. Conducting internal surveys and a focus group discussion can also help reveal much about the new intervention’s potential for success. After these practical internal sessions, the material and select sections of the intervention are pilot tested for understanding and applicability with select community members (selected because they are a good representation of the population of focus). The goal is to ensure they too can identify with the content and format. The feedback can then be used to put the final “touches” to the intervention for pilot launching.

#### Developing and Testing Assessment/ Evaluation Tools

To assess intervention outcomes both written and biometric screening tools (pre/post-surveys collecting demographic information and assessing behaviors, collection of biometric and laboratory measurements) are recommended. Plans should be made to compensate participants (e.g., gift cards to a frequently used grocery store) for the time spent answering questions.

#### Written Assessments

Administrating written assessments to a group of individuals with a wide range of literacy levels can be challenging. Limiting the number of questions and careful selection of their sequence to minimize fatigue and improve response accuracy is critical. Written assessment tools, even validated ones, containing Likert-scaled questions may be confusing and perceived as tricky and repetitive. Therefore, adjustments may be necessary, keeping in mind that participants’ cultural background could affect their response pattern (Wang, et al., 2008). Lastly, it is best to not use subscript numbers (used to facilitate data entry) and other potentially distracting characters.

##### Biometric Assessments

Similar to written assessments, interpretation of standard measurements may need to be adapted based on ethnicity or race - e.g., hemoglobin A1C for Latinos, waist measurements for Africans, Asians and Native Americans (Cavagnolli, 2017; Klimentidis et al., 2016).

##### Qualitative data

Focus group discussion questions and recording devices should also be prepared for use immediately following the intervention for a more comprehensive assessment.

### Step 4: Pilot Test and Evaluate Intervention

#### Implementing New Intervention in Pilot Groups

Once the intervention and assessment tools have been created, CHWs should plan to pilot the intervention at different locations and in different settings, using a variety of formats and time frames. This allows the team to determine what works best.

#### Evaluating Process, Outcomes and Population Fit

To evaluate the process, it is best to have a third party observe and document program delivery to assure fidelity and reception. Field notes from CHWs facilitating the sessions can also add important perspectives. Besides this process evaluation, assessing outcomes and best population fit can be done by asking participants to evaluate the setting, material and program in general.

### Step 5: Revise Program

#### Re-assessing Process, Outcomes and Sustainability

During this step the team meets to analyze the process and outcomes as expressed by participants in focus group discussions and surveys, in CHWs field notes and by the third-party evaluator. The goal is to answer the following questions: Is the material/intervention being implemented as planned? Is the material and intervention clearly understood independently of literacy level? If not, what adjustments are needed to better accommodate this priority population?

#### Revising the Content and Format of the New Intervention

Practical steps are taken to adjust program implementation, content and materials according to feedback gathered and received during the program revision.

### Step 6: Launch Intervention

This step is self-explanatory for the most part. Recruitment may consist of creating appropriate flyers and having CHWs advertise the intervention at health fairs, community centers and churches. After gathering names, addresses and telephone numbers of participants and then calling them to see which locations and time frames work best for the majority, a starting date and time is set. The timing of weekly sessions should take into consideration transportation options, participants’ responsibilities (e.g., meal preparation), mealtimes, holidays, children’s school schedule, childcare and financial limitations. In selecting locations to hold the meetings, community centers or church buildings participants are familiar with usually work best.

#### Analyses

The process of developing this model occurred while creating and implementing a weight management/diabetes prevention intervention among low-income monolingual (Spanish-speaking only) Latinas with limited literacy living in Southern California - the state with the lowest literacy level in the nation (World Populations Review, 2024). This population also happens to have one of the highest rates of obesity (Powell-Wiley et al., 2021) and have characteristics of the five largest groups identified with low literacy (Kutner, 2006; Rikard,

2016). Notes about the process and the rationale for selected choices were carefully kept by the research team. A summary is shared below.

#### Practical application of Step 1

While interacting informally with Latino community members at community centers and local clinics the researchers gathered information about values associated with obesity and potential barriers to a weight management program, then engaged interviewed individuals on the topic of obesity. Each participant received a gift card as a “Thank you” gift to compensate for their time.

Then, after completing a literature review, the two researchers conducted two focus group discussions (FGDs) with community health workers (N=15) from the community. All FGD participants were asked to read and sign an informed consent form in their preferred language which had been reviewed and approved by the oversight institution’s Institutional Review Board. To protect confidentiality, transcripts were de-identified, using 2017 computer software program MaxQDA® (v.12, Berlin, GDR) (Verbi, 2015).

In the process, the researchers confirmed that several previously published concepts associated with obesity existed. The main reasons cited for obesity in this population were ignorance, lifestyle habits, food availability, family and cultural values, and immigration into the USA. Other reasons given included media, genes, stress, overall health, and lack of time.

Creating a visual representation of the key issues at stake helped to gain a clearer perspective on the topic (see Figure 2). Some perceived needs included access to healthy food options, easier (easy-to-understand and apply) ways of determining quantity and types of recommended foods, available time to attend classes and cook healthy meals, viable alternatives to walking in their neighborhoods (preferred option for physical activity) and stress management skills.

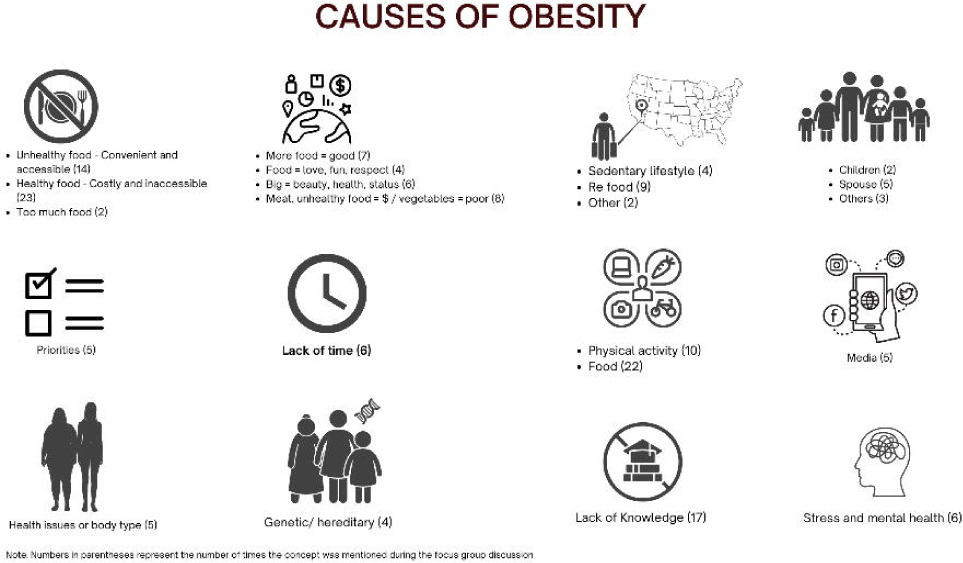

Barriers to adopting healthy behaviors were also identified: These included *familismo* (a “strong orientation and commitment toward the family”), the role of women (Davila, 2011; Dacosta, 2023), the belief that obesity and eating meat are evidence of prosperity (in contrast to beans, vegetables, and weight loss). Still, even within the local Latino population some values differed by age, country of origin and acculturation level. Thus, it became obvious that these issues would need to be carefully and tactfully addressed.

#### Practical application of Step 2

The interdisciplinary team consisted of five community health workers (one of which was obese and had recently been diagnosed with prediabetes), two physicians (one being the principal investigator), three medical students and a nutritionist/social epidemiologist. The input of all these members, especially during the initial two weeks of brainstorming/program creation, assured medical accuracy as well as insights and scientific information from a nutritionist and healthcare provider perspective. To encourage honest and “non-hierarchical” communication and avoid intimidation, academic and professional accomplishments of fellow team members were not revealed for the first two weeks. Medical students were also told to not wear clothing that identified them as medical students or physicians. Thus, CHWs were not aware that they were interacting with medical students or scientists.

The team identified an intervention framework that used proportions instead of portion sizes, had an easily identifiable color-coded list of “recommended foods,” required no calorie counting and no reading of food labels (Seale et al., 2010). Recognizing that many Latino women with low income already lead high stress lives even before dealing with the stress of adopting new behaviors, the team chose to supplement that framework with simple easy stress management techniques from the Community Resiliency Model (CRM), a model adaptable for lay individuals which has been found to be effective cross culturally (Freeman, 2022; Trauma Resource Institute, n.d.). To facilitate adult/popular education style the content of the hands-on applications was also divided into adult teachable units throughout the program and the sessions were held with everyone sitting in a circle.

Nutrition and scientific content (e.g., dietary fiber, brain changes associated with stress, sugar-sweetened beverages) were first to be presented with clear easy explanations to which more complex concepts were added. Interactive components included analyzing and comparing local restaurants menus, grocery shopping and a cooking class. After selecting ingredients of “traditional typical” recipes to favorite dishes, the team shopped at a local store for ingredients to prepare these dishes, as well as alternative healthy variations. Then, as part of the cooking class, meals were prepared, rated by all and ranked according to taste, cultural acceptability, and appearance as well as likelihood of being used in the future. In keeping with *familismo,* tips to “sell” the recommended foods to family members, such as using creative positive rather than restrictive terms, were also shared.

The didactic material and participants’ workbooks were created following recommended guidelines for written communication to be used among individuals with limited literacy. We used photographs of local individuals that participants could identify with and selected colors compatible with the local Latino culture. Figure 3 shows examples of changes made to the original material. Instead of providing participants with the entire course material, each week’s lesson would be distributed weekly in the form of loose sheets that fit in a three-ring binder. Lastly, given the potential limitations of program delivery sites, easy-to-carry flip charts were created for facilitators’ use.

**Figure 3.**
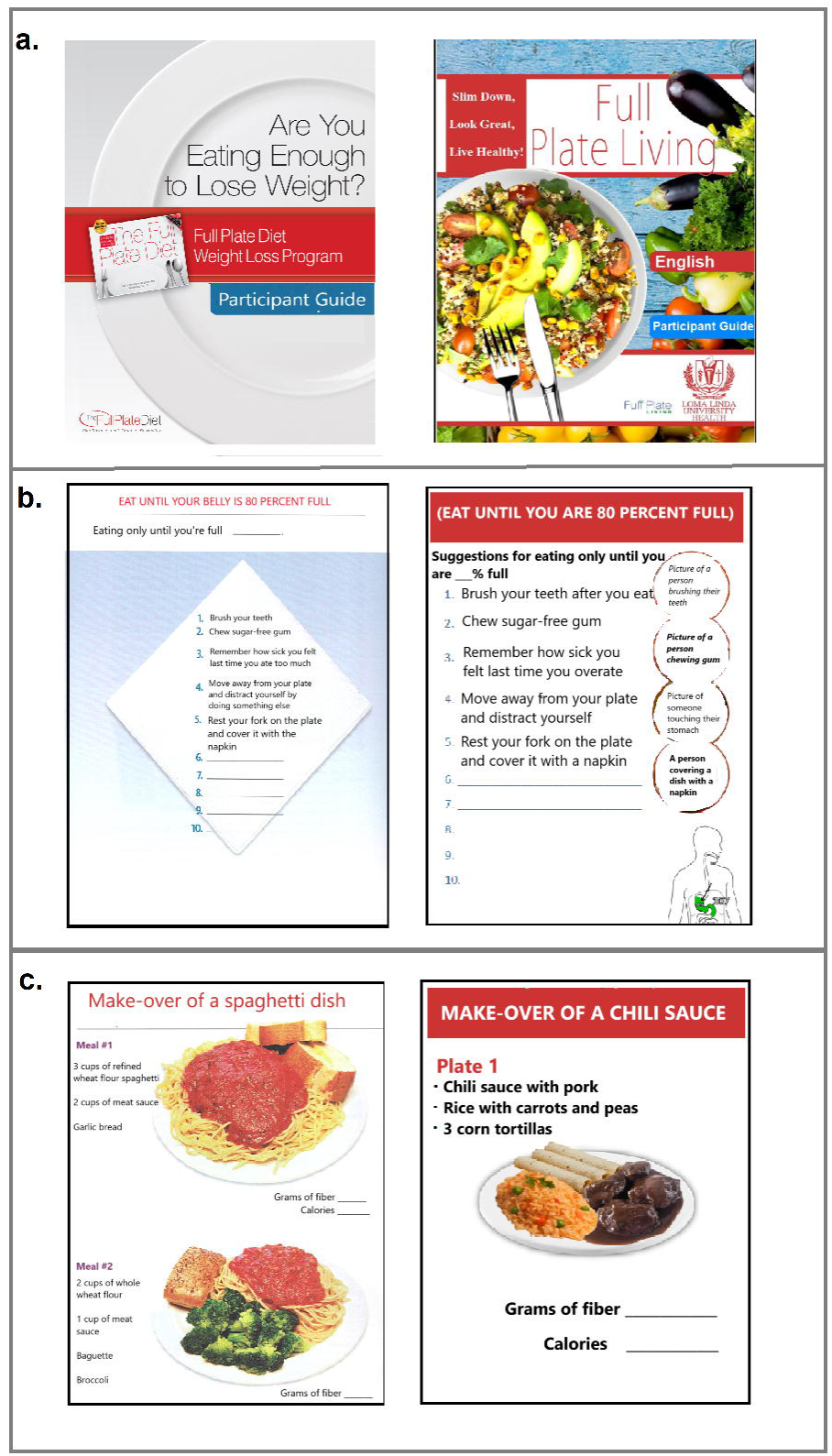
Examples of adaptations made to the material (left column are pictures/pages from the original book, right column are adapted pages) **a.** (left) cover page: few colors and mostly words format: wire-bound (right) cover page: relatable graphics and colors compatible with the Latino culture format: loose pages (for use with 3-ring binder) **b.** (left) wording: small font and more words picture: minimal and not clearly illustrative of concepts conveyed on the page (right) wording: larger font, less words with contrasting background picture: several pictures clearly illustrating concepts mentioned on the page Note: page title was translated from Japanese "Hara Hachi Bu" **c.** (left) more words, small font, less "white space" and 2 variations of a typical American meal (right) less words, larger font, more "white space" and only 1 typical Mexican dish

CHW team members took turns conveying the key messages in an easy-to-understand culturally appropriate and relevant way. They practiced facilitating the sessions until they felt that the intervention would be relatable and easily understood. During this process, all team members also applied program recommendations in their own lives and with family members. Thus, they could attest to the success of the program and identify future participants’ possible challenges.

#### Practical application of Step 3

Pretesting of the material was done by the same diverse team with CHWs giving feedback about the appropriateness of each of the pictures and pages and selecting the final material. The last session of the intervention was spent implementing the concepts learned in the preparation of an adapted version of traditional favorite dishes. A cooking demonstration for guests and co-workers was held at the end.

Because of the characteristics of this group - and based on studies among similar populations – in addition to the usual demographic questions (education level, income, and health insurance), survey questions also assessed household size, neighborhood type, transportation, as well as access to healthy foods. Easier questions were placed towards the end of two separate surveys to be answered at different times. Besides limiting the number of questions and to minimize fatigue and improve response accuracy we determined that it was best to split them into multiple shorter surveys and to move easier questions towards the end. Because social determinants of health can dramatically affect attendance to programs and healthcare, questions on these should be included early on. A decision was made about the type of gift card (to a store commonly used by participants) to be given in exchange for answering the surveys.

We would also incorporate a “teaching moment” session during the post-survey for health counseling of participants, a rare opportunity for individuals to gain relevant information about their health. Team members participated in a focus group discussion and answered pre and post-surveys.

#### Practical application of Step 4

The program was piloted at three different locations and settings (a church building, a community center, a gathering center for food pantry recipients) by two community health workers each. Participants in the three pilot groups (N=46) were Latinos, most were mono-lingual Spanish speakers with low acculturation who were married or living with a partner. All 3 groups had comparable socio-economic status profiles. While we used the same materials - program lengths, delivery methods, and frequency of sessions varied: one was held twice a week and the others only once a week. Sessions lasted 2 hours on average and We also had meetings in both morning and evenings to accommodate participants’ schedules.

While most participants decided to complete the surveys by themselves (following instructions from a CHW using flip chart prompts), some needed a CHW to discreetly help them with reading and answering questions. To help participants develop new healthy behaviors, water and culturally appropriate snacks were provided at each session. Interactive activities included food exchange, a grocery shopping trip and a cooking class as described above. The cost of traditional and of adapted meals were compared, demonstrating that cost need not be a major barrier to implementing the intervention.

Small surprise gifts and weekly delivery of didactic material acted as teasers to return and helped increase retention. Lastly, to promote program sustainability, the team decided to encourage participants to create small support groups within the larger group. While classes were delivered by CHWs, occasional “ask the expert” surprise segments by healthcare professionals were interspersed throughout the intervention, an opportunity to provide answers to questions that exceeded CHW training. In addition to the group work, participants had the option to discuss specific barriers and sensitive issues privately with a CHW, either in person or by telephone. CHWs called participants weekly to check on their progress and to remind them of the next class.

To maintain contact with participants and follow-up on participants after the intervention, CHWs continued with high levels of interaction (phone calls, role playing and reminders) resulting in the realization that 70% participant were still implementing some of the learned practices over the course of one year even though the intervention only lasted 3 months. This happened despite the COVID-19 pandemic (*citation removed for masked peer-review)*.

Outcomes measures included weight and changes in eating behavior. In addition to having a person observe program delivery to ensure fidelity and reception, we asked participants to share their feedback. A focus group discussion was conducted at program completion and a celebratory graduation was held with family members in attendance.

Since we deliberately varied program characteristics, we were able to determine which modality worked best for our target groups: participants preferred the use of flip charts and the ten-week option (vs biweekly meetings for five weeks). We also discovered which one had the highest program completion and retention rate. While outcome results were similar across all three groups, the time of day was not that important as long as it accommodated the group.

#### Practical application of Step 5

The team met on several occasions to reassess the program. To improve results, we realized we needed to allocate more time for assessments and feedback and needed to revise the material.

To help individuals set more accurate weight loss goals we added an orientation session and additional time during the second session to discuss lab results with the group.

Material revision included reducing the number of words, highlighting some key words, adding more pictures, more “white space” (empty space in a page) and more contrasting colors. Competitions between groups would be added to motivate participants: inexpensive pedometers would be given (since many didn’t have smart phones) as a reward for physical activity, biometric changes and to those who encouraged other group members.

To increase engagement and promote sustainability, we also needed to motivate individuals to connect outside of session times and select accountability partners within their small support group so as to provide each other with more social support. If necessary, and based on group dynamics, CHWs would help each group identify and select “leaders” to lead on a monthly rotation basis. A private Facebook page and a “WhatsApp” group were created so individuals could showcase their new dishes, resources, and progress with one another. Lastly, we realized that it made sense culturally and for better group dynamics to keep the groups only to women because they requested that men attend in separate groups (men tended to dominate the conversations and dismiss women’s opinions).

Regarding assessment of food intake (for our outcome study), a separate flipchart was created showing pictures of food categories (vegetables, fruits, legumes, beverages, desserts….) available in their neighborhoods and eaten by most community members. It would be used as a reference to help participants accurately identify and report the types of foods they had been consuming.

## Results

Sixteen months after starting the process (including the delivery of the pilot studies), the team was finally ready to officially launch the intervention on a larger scale. The CHWs went to community centers, health fairs and churches to advertise the program, using flyers that had been created jointly. Other participants (including family members and acquaintances of CHWs) joined through word of mouth. We were especially careful in the selection of meeting locations (trust and safety mattered as we were in a period of immigration raids).

Approximately one month later, the first meeting of the intervention was held. To date the program has been implemented successfully among more than 200 participants (after the above pilot testing) and resulted in comparable outcomes in both food secure and food insecure participants (*citation removed for masked peer review*). The reading level also turned out to be at 4^th^ grade level using the Flesch Kincaid method (Flesch, 1948). While the 3 months data are strong and showed success among different levels of food insecurity, the long-term (1 year) data showed retention was high especially among those meeting in the same space (manuscript submitted for publication). Of note, more than a year after program completion, these participants continue to share tips with each other on social media.

We believe that the LACASA principles can be applied even when program restrictions such as social distancing are required as we have successfully used them in an online format during the pandemic (American Public Health Association Annual Conference, 2022).

### COVID-19 pandemic revision

The COVID-19 pandemic led us to ask and answer questions which led to a major program revision: needs, program delivery, didactic material, timing all changed. To maintain social distancing, we had to move most of the program online (Zoom) and online groups were reduced to a maximum of 10 participants. A few (mostly those with low comfort with/access to technology) chose to attend in person while maintaining social distancing. Flipchart content was transformed into power point presentations, some with videos, and interactive games (e.g., “Russian Roulette” game) were included to help retain participants’ attention and motivation. Key points for each session were reduced and Zoom breakout rooms became the main platform for participant-to-participant interaction. Due to the severe impact of the pandemic on mental health more emphasis was placed on the mental health component of the program. Small prizes gained during the sessions – except for gardening kits and garden starters - were retained until program completion and given at the “graduation ceremony”. During the holiday season (an emotionally difficult season during the first year of the pandemic), we also created a drop box near the home of one of the CHWs where participants could leave anonymous encouraging messages for one another. A digital scale was also left nearby for those who did not own a scale and wanted to keep track of their weight.

Including questions about the connection between fellow-participants may indicate how much social connection occurs within each group/setting, an important factor for long-term sustainability. Paper surveys were replaced with Qualtrics or telephone surveys and included questions assessing participants’ comfort with technology. Overall, more staff time was now required (to provide support with technology, to prepare and teach each session, follow-up on participants, help participants answer surveys, allow each participant to maintain social distancing during assessments, etc…). Additional equipment (personal protective equipment) and space were also necessary. We found that participants still benefited from the program, although results varied based on modality (attending in a group or alone, online vs in-person) (American Public Health Association, 2022).

## Discussion and Lessons Learned

There is no doubt that producing an effective sustainable program for low-literacy, low-income minorities or cultural subgroups requires time. Throughout the implementation, the lower the reading comprehension and the more social determinant needs, the more time was required. We learned that CHWs must allow additional time to provide resources and emotional support and help participants problem-solve, and to “translate” written material, especially assessments, into practical terms.

Although it is possible for the program development to take less time, especially if more funding is available, the entire process of adapting an existing traditional approach to create our LACASA intervention took more than a year. For example, even with materials created for low literacy/numeracy levels one should expect some individuals to require individual attention to understand the material content.

This is similar to what has been reported by Seligman et al (2007) and Rosas et al. (2018) who also spent more than a year creating a program. This may seem too costly an initial investment, but the short and long-term results are worthwhile, considering our successful retention of participants over more than one year despite COVID-19 challenges [66] and the cost of low or limited literacy and chronic diseases (Chapel et al., 2017; Visscher et al., 2018).

With so many obstacles, low-literacy, low-income individuals face before starting a lifestyle or weight loss program, each program should include assessing the environmental social determinants of health of their priority population. It is important to make sure that individuals have access to all they need to apply what they are learning: healthy, fresh food, appropriate timing of classes, childcare, places to exercise, and reliable access to the internet (if attending online). Encouraging participants to find a way to eat the recommended food can be challenging for individuals depending on food banks which provide less than healthy options or where the only local stores are liquor stores. However, this cannot be assumed to be the case for every community or group. In our experience, using a low-literacy approach to a weight management program, incorporating culturally acceptable healthy dishes, and providing resources to maximize physical activity, stress management and healthy food and beverage consumption resulted in a program that was easy, simple, and truly “speaks” to the community it was intended to reach.

Participants enrolled in our LACASA program with low-income and low-literacy levels appreciated having access to more appropriate material and community health workers to explain and teach them about critically important preventive health behaviors. This helped them make lifestyle changes that - they now understand - are critical to their health, without undermining their cultural roots.

We found that our word parsimonious material was welcomed independently of the literacy level of our participants. This is similar to what was concluded by another study which reported that creating material understandable to individuals with low literacy does not necessarily turn off those with higher literacy level (Weiss, 1998). Participants reported high acceptability and appropriateness. The focus groups suggested that most content was understood and applied in ways Although we would argue that it is not necessary to start from scratch every time, every community has its own idiosyncrasies. We feel that for similar cultural sub-groups - and these could include English speakers and American-born individuals – gathering some early feedback to the approach to assure relevance of the material and allow for minor adaptations will suffice. Language preference and acculturation vary. So, it is important to have at least a bilingual CHW.

We also feel that leadership and co-ownership of our CHWs was just as important as the material, especially as we discovered that certain misunderstandings remain despite years of exposure to new health information (Easton et al., 2013; Trettin & Musham, 2000). Their ability to relate and connect with their audience makes them the ideal and trusted voices to clarify issues and if needed, provide a counter narrative. Furthermore, the respectful (and moderate) inclusion of traditional health professionals willing to work with the CHWs and the community with cultural humility was also critically important. Their involvement was welcome and well received by participants, a response also reported elsewhere (Brown et al., 2007). Lastly, inadvertently, CHWs developed a support group and held each other accountable as good role models to their communities. The interdisciplinary team that brainstormed together also remained close.

### Strengths and Limitations

One weakness of this study is that the initial team involved a small number of CHWs (N=5) and only 3 pilot groups totaling 46 participants to develop it. However, we collected rigorous formative, process, quantitative outcome and contextual qualitative data across all phases. Also, the principles of the program have not been tested in other populations and funding was limited (which impacted the duration of some steps). Thus, it may not be fully replicable without further adaptations or major changes. Because we decided to only explore feasibility and effectiveness among women in the revised intervention, we have no male comparison group. The intervention was not tested against an evidence-based intervention (i.e., NDPP Latino version) using random assignment, all currently considered as next steps. However, we were able to successfully apply the model to create an online adaptation during the pandemic, thus demonstrating its relevance and applicability during challenging times and major population crises.

### Implications for Theory, Policy and / or Practice

Partnership between researchers, health educators, physicians, medical students and CHWs using a community-participatory approach to create interventions has the potential of providing a culturally relevant lifestyle program that could be used to reach high-risk, low-income underserved populations and bring much needed education and health benefits to these communities.

In view of the increasingly high percentage of the US population with limited literacy, it is important to reconsider the use and content of written material, especially when dealing with low-income vulnerable populations, and to invest in programs that are a better fit. This entails carefully listening to the population of interest and allowing adequate time to “translate” programs. Investing in CHWs for this purpose seems to provide the best return on investment and bodes well for the sustainability of such programs. Furthermore, we believe this framework could be helpful to those wishing to conduct participatory action research and incorporate similar strategies as those used in this study.

With the lingering threat of COVID-19 remaining, online program options promise to stay popular. As we had to move all program delivery on-line due to the COVID-19 pandemic, we believe that careful application of the LACASA model, using small group delivery on-line is possible and sustainable, especially as more participants have access to smart phones, although this may require even more time and creativity.

We hope that our sharing of this model process used, and of the lessons learned will serve as a valuable resource to help guide and accelerate the development of other lifestyle-based interventions led by community health workers for individuals challenged with functional literacy living in resource-constrained settings, independently of their culture or subculture.

## Data Availability

The data produced are not available.

## Acknowledgements

We would like to express our thanks to El Sol Neighborhood and Educational center for supporting our community health workers. We are also grateful for the hard work of research assistants Marisol Lara and Raveena Chara, community health workers Vilma Lopez, Silvia Guerra, Maria Elena Cháirez and Marisa Aguero, medical students (now physicians) Shevel DaCosta Davis, Simone Deshields, Lauren Miller, and Dr. Camille Clarke for their critical input during the first phase of curriculum development.

## References

American Anti-Slavery Committee. (n.d.). Slavery and the International Slave Trade in the United States of America. American Anti-Slavery Committee | The Gilder Lehrman Center for the Study of Slavery, Resistance, and Abolition (yale.edu)

American Public Health Association. (2022). APHA 2022 Annual meeting and Expo. https://apha.confex.com/apha/2022/meetingapp.cgi/Paper/517970

Association of clinicians for the underserved/patient education material (2020). *General Diabetes Patient Education Handouts*. http://clinicians.org/our-issues/acu-diabetes-patient-education-series/

Baig, A. A., Locklin, C. A., Wilkes, A. E., Oborski, D. D., Acevedo, J. C., Gorawara-Bhat, R., Quinn, M. T., Burnet, D. L., & Chin, M. H. (2012). "One Can Learn From Other People’s Experiences": Latino adults’ preferences for peer-based diabetes interventions. The Diabetes educator, 38(5), 733–741.

Bekteshi V. (2024). Decoding acculturative stress and psychological distress in Mexican immigrant women: insights from a path mediation analysis. BMC women’s health, 24(1), 667. 10.1186/s12905-024-03494-1

Bos-Touwen, I., Jonkman, N., Westland, H., Schuurmans, M., Rutten, F., de Wit, N., & Trappenburg, J. (2015). Tailoring of self-management interventions in patients with heart failure. Current heart failure reports, 12(3), 223–235. 10.1007/s11897-015-0259-3

Boutemen, Miller. (2023). Readability of publicly available mental health information: A systematic review. Patient Education and Counseling, 111(107682). 10.1016/j.pec.2023.107682.

Brown, C., Hennings, J., Caress, A. L., & Partridge, M. R. (2007). Lay educators in asthma self management: Reflections on their training and experiences. Patient Education and Counseling, 68(2), 131–138. 10.1016/j.pec.2007.05.009

Burki, T. K. (2020). El susto and the Mexican sugar tax. The Lancet Diabetes & Endocrinology, 8(4), 277. doi: 10.1016/S2213-8587(20)30065-6

Cavagnolli, G., Pimentel, A.L., Freitas, P.A., Gross, J.L., & Camargo, J.L. (2017). Effect of ethnicity on HbA1c levels in individuals without diabetes: Systematic review and meta-analysis. PLoS One, 12(2):e0171315. doi:10.1371/journal.pone.0171315

Centers for Disease Control and Prevention. (2018). *Logic Models – CDC approach to evaluation.* Logic Models - Program Evaluation - CDC

Centers for Disease Control and Prevention. (2023). Understanding health literacy. https://www.cdc.gov/healthliteracy/learn/understanding.html#Arelimitedhealthliteracyandlimitedliteracythesameproblem

Chapel, J.M., Ritchey, M.D., Zhang, D., & Wang, G. (2017). Prevalence and Medical Costs of Chronic Diseases Among Adult Medicaid Beneficiaries. American Journal of Preventive Medicine,53(6S2):S143-S154. doi:10.1016/j.amepre.2017.07.019

Color meaning. (n.d.) *Color symbolism in different cultures around the world*. https://www.color-meanings.com/color-symbolism-different-cultures/

CommunicateHealth, Inc. (2023, March 9). A Framework for Equity-Centered Health Communication. https://communicatehealth.com/wp-content/uploads/ch-echc-framework.pdf

Da Costa M. (2023). How Culture Impacts Health: The Hispanic Narrative. Creative nursing, 29(3), 273–280. 10.1177/10784535231211695

Davila, Y. R., Reifsnider, E., & Pecina, I. (2011). Familismo: influence on Hispanic health behaviors. Applied nursing research, 24(4), e67–e72. 10.1016/j.apnr.2009.12.003

De Bortoli, M., & Maroto, J. (2001). Colours Across Cultures: Translating Colours in Interactive Marketing Communication. http://globalpropaganda.com/articles/TranslatingColours.pdf

Easton, P., Entwistle, V. A., & Williams, B. (2013). How the stigma of low literacy can impair patient-professional spoken interactions and affect health: insights from a qualitative investigation. BMC health services research, 13, 319. 10.1186/1472-6963-13-319

ElSayed, N. A., Aleppo, G., Aroda, V. R., Bannuru, R. R., Brown, F. M., Bruemmer, D., Collins, B. S., Hilliard, M. E., Isaacs, D., Johnson, E. L., Kahan, S., Khunti, K., Leon, J., Lyons, S. K., Perry, M. L., Prahalad, P., Pratley, R. E., Seley, J. J., Stanton, R. C., …Young-Hyman, D. on behalf of the American Diabetes Association (2023). 5. Facilitating Positive Health Behaviors and Well-being to Improve Health Outcomes: Standards of Care in Diabetes-2023. Diabetes care, 46(Supple 1), S68–S96. 10.2337/dc23-S005

Flesch, R. (1948). A new readability yardstick. Journal of Applied Psychology, 32(3), 221–233. 10.1037/h0057532

Freeman, K., Baek, K., Ngo, M., Kelley. V., Karas, E., Citron S., & Montgomery, S. (2022). Exploring the Usability of a Community Resiliency Model Approach in a High Need/Low Resourced Traumatized Community. Community Mental Health Journal, 58 (679–688). 10.1007/s10597-021-00872-z

Gajjar, A. A., Patel, S., Patel, S. V., Goyal, A., Sioutas, G. S., Gamel, K. L., Salem, M. M., Srinivasan, V. M., Jankowitz, B. T., & Burkhardt, J. K. (2024). Readability of cerebrovascular diseases online educational material from major cerebrovascular organizations. Journal of neurointerventional surgery, jnis-2023–021205. Advance online publication. 10.1136/jnis-2023-021205

Grant, M., Wilford, A., Haskins, L., Phakathi, S., Mntambo, N., & Horwood, C. M. (2017). Trust of community health workers influences the acceptance of community-based maternal and child health services. African journal of primary health care & family medicine, 9(1), e1–e8. 10.4102/phcfm.v9i1.1281

Han, A., & Carayannopoulos, A.G. (2020). Readability of Patient Education Materials in Physical Medicine and Rehabilitation (PM&R): A Comparative Cross-Sectional Study. Physical Medicine and Rehabilitation, 12(4):368–373. doi:10.1002/pmrj.12230

Handtke, O., Schilgen, B., & Mösko, M. (2019). Culturally competent healthcare - A scoping review of strategies implemented in healthcare organizations and a model of culturally competent healthcare provision. PloS one, 14(7), e0219971. 10.1371/journal.pone.0219971

Interaction Design Foundation (2021, November 4). *What is Color Symbolism?* https://www.interaction-design.org/literature/topics/color-symbolism

Jacob, V., Chattopadhyay, S. K., Hopkins, D. P., Reynolds, J. A., Xiong, K. Z., Jones, C. D., Rodriguez, B. J., Proia, K. K., Pronk, N. P., Clymer, J. M., & Goetzel, R. Z. (2019). Economics of Community Health Workers for Chronic Disease: Findings From Community Guide Systematic Reviews. American journal of preventive medicine, 56(3), e95–e106. 10.1016/j.amepre.2018.10.009

Kennedy, B. M., Rehman, M., Johnson, W. D., Magee, M. B., Leonard, R., & Katzmarzyk, P. T. (2017). Healthcare Providers versus Patients’ Understanding of Health Beliefs and Values. Patient experience journal, 4(3), 29–37

Klimentidis, Y. C., Arora, A., Zhou, J., Kittles, R., & Allison, D. B. (2016). The Genetic Contribution of West-African Ancestry to Protection against Central Obesity in African-American Men but Not Women: Results from the ARIC and MESA Studies. Frontiers in genetics, 7, 89. 10.3389/fgene.2016.00089

Knowles, M. S. (1968). Andragogy, not pedagogy. Adult Learning, 16(10), 350–352.

Kreuter, Matthew W., Susan N. Lukwago, Dawn C. Bucholtz, Eddie M. Clark, and Vetta Sanders-Thompson. 2003. “Achieving Cultural Appropriateness in Health Promotion Programs: Targeted and Tailored Approaches.” Health Education & Behavior 30 (2): 133–46. 10.1177/1090198102251021.

Lee, D. M., Grose, E., & Cross, K. (2022). Internet-based patient education materials regarding diabetic foot ulcers: readability and quality assessment. Journal of Medical Internet Research Diabetes, 7(1), e27221.

Lipari, M., Berlie, H., Saleh, Y., Hang, P., & Moser, L. (2019). Understandability, actionability, and readability of online patient education materials about diabetes mellitus. American journal of health-system pharmacy, 76(3), 182–186. 10.1093/ajhp/zxy021

Literacy Pittsburgh. (2024). *Challenges of low literacy*. The Challenge | Causes of Low Literacy |Literacy Pittsburgh

National Literacy Institute. (n.d.). Literacy Statistics 2024-2025 (Where are we now). Literacy Statistics | National Literacy (thenationalliteracyinstitute.com)

López, L., Tan-McGrory, A., Horner, G., & Betancourt, J. R. (2016). Eliminating disparities among Latinos with type 2 diabetes: Effective eHealth strategies. Journal of diabetes and its complications, 30(3), 554–560. 10.1016/j.jdiacomp.2015.12.003

Lueken, A., Mangan, M., & Smaellie, S. (2022). The Hand that Rocks the Cradle Cannot Read this Title: The Multi-Generational Effect of Illiteracy in the Lives of Black American Women. Utah Women’s Health Review. doi: 10.26054/0d-86nh-yyqh

Lyons, B., & Dolezal, L. (2024). Shame, health literacy and consent. Clinical ethics, 19(2), 150–156. 10.1177/14777509231218203

Martin, L. T., Schonlau, M., Haas, A., Derose, K. P., Rudd, R., Loucks, E. B., Rosenfeld, L., & Buka, S. L. (2011). Literacy skills and calculated 10-year risk of coronary heart disease. Journal of general internal medicine, 26(1), 45–50. 10.1007/s11606-010-1488-5

Menendian, S., Gambhir, S., & Gailes, A. (2021, June 21). The Roots of Structural Racism Project. The Roots of Structural Racism Project | Othering & Belonging Institute (berkeley.edu)

Moffett, M. L., Kaufman, A., & Bazemore, A. (2018). Community Health Workers Bring Cost Savings to Patient-Centered Medical Homes. Journal of community health, 43(1), 1–3. 10.1007/s10900-017-0403-y

Nash, E., Bickerstaff, M., Chetwynd, A. J., Hawcutt, D. B., & Oni, L. (2023). The readability of parent information leaflets in paediatric studies. Pediatric research, 94(3), 1166–1171. 10.1038/s41390-023-02608-z

New York University Health Sciences Library. (2023). *Health Literacy and Patient Education*. Toolkits and Guides - Health Literacy and Patient Education - Subject Guides at NYU HealthSciences Library

Novin, S. A., Huh, E. H., Bange, M. G., Hui, F. K., & Yi, P. H. (2019). Readability of Spanish-Language Patient Education Materials From RadiologyInfo.org. Journal of the American College of Radiology : JACR, 16(8), 1108–1113. 10.1016/j.jacr.2018.12.036

Painter, S. L., Ahmed, R., Hill, J. O., Kushner, R. F., Lindquist, R., Brunning, S., & Margulies, A. (2017). What Matters in Weight Loss? An In-Depth Analysis of Self-Monitoring. Journal of medical Internet research, 19(5), e160. 10.2196/jmir.7457

Parikh, N. S., Parker, R. M., Nurss, J. R., Baker, D. W., & Williams, M. V. (1996). Shame and health literacy: the unspoken connection. Patient education and counseling, 27(1), 33–39. 10.1016/0738-3991(95)00787-3

Pignone, M., DeWalt, D. A., Sheridan, S., Berkman, N., & Lohr, K. N. (2005). Interventions to improve health outcomes for patients with low literacy. A systematic review. Journal of general internal medicine, 20(2), 185–192. 10.1111/j.1525-1497.2005.40208.x

Powell-Wiley, T. M., Poirier, P., Burke, L. E., Després, J. P., Gordon-Larsen, P., Lavie, C. J., Lear, S. A., Ndumele, C. E., Neeland, I. J., Sanders, P., St-Onge, M. P., & American Heart Association Council on Lifestyle and Cardiometabolic Health; Council on Cardiovascular and Stroke Nursing; Council on Clinical Cardiology; Council on Epidemiology and Prevention; and Stroke Council (2021). Obesity and Cardiovascular Disease: A Scientific Statement From the American Heart Association. Circulation, 143(21), e984–e1010. 10.1161/CIR.0000000000000973

Price, J. H., Khubchandani, J., & Webb, F. J. (2018). Poverty and Health Disparities: What Can Public Health Professionals Do?. Health promotion practice, 19(2), 170–174. 10.1177/1524839918755143

Racoma, B. (2019, July 26). Day Translations Blog. Color symnbolism – Psychology across Cultures. Color Symbolism and Psychology Across Different Cultures (daytranslations.com)

Ridley, M., Rao, G., Schilbach, F., & Patel, V. (2020). Poverty, depression, and anxiety: Causal evidence and mechanisms. *Science (New York*, N.Y.), 370(6522), eaay0214. 10.1126/science.aay0214

Rothman, R. L., DeWalt, D. A., Malone, R., Bryant, B., Shintani, A., Crigler, B., Weinberger, M., & Pignone, M. (2004). Influence of patient literacy on the effectiveness of a primary care-based diabetes disease management program. JAMA, 292(14), 1711–1716. 10.1001/jama.292.14.1711

Rosas, L. G., Lv, N., Lewis, M. A., Venditti, E. M., Zavella, P., Luna, V., & Ma, J. (2018). A Latino Patient-Centered, Evidence-Based Approach to Diabetes Prevention. Journal of the American Board of Family Medicine, 31(3), 364–374. 10.3122/jabfm.2018.03.170280

Sanders, L. M., Perrin, E. M., Yin, H. S., Bronaugh, A., Rothman, R. L., & Greenlight Study Team (2014). "Greenlight study": a controlled trial of low-literacy, early childhood obesity prevention. Pediatrics, 133(6), e1724–e1737. 10.1542/peds.2013-3867

Sasson, C., Haukoos, J. S., Ben-Youssef, L., Ramirez, L., Bull, S., Eigel, B., Magid, D. J., & Padilla, R. (2015). Barriers to calling 911 and learning and performing cardiopulmonary resuscitation for residents of primarily Latino, high-risk neighborhoods in Denver, Colorado. Annals of emergency medicine, 65(5), 545–552.e2. 10.1016/j.annemergmed.2014.10.028

Seale, S.A., Sherard, T., & Flemming, D. (2010). The Full Plate Diet. Bard Press.

Seligman, H. K., Wallace, A. S., DeWalt, D. A., Schillinger, D., Arnold, C. L., Shilliday, B. B., Delgadillo, A., Bengal, N., & Davis, T. C. (2007). Facilitating behavior change with low-literacy patient education materials. American journal of health behavior, 31 *Suppl 1*(0 1), S69–S78. 10.5555/ajhb.2007.31.supp.S69

Shelton, R. C., Goldman, R. E., Emmons, K. M., Sorensen, G., & Allen, J. D. (2011). An investigation into the social context of low-income, urban Black and Latina women: implications for adherence to recommended health behaviors. Health education & behavior : the official publication of the Society for Public Health Education, 38(5), 471–481. 10.1177/1090198110382502

Singh, S. P., Qureshi, F. M., Borthwick, K. G., Singh, S., Menon, S., & Barthel, B. (2022). Comprehension Profile of Patient Education Materials in Endocrine Care. Kansas journal of medicine, 15, 247–252. 10.17161/kjm.vol15.16529

Stormacq, Coraline, Stephan Van den Broucke, and Jacqueline Wosinski. 2019. “Does Health Literacy Mediate the Relationship between Socioeconomic Status and Health Disparities? Integrative Review.” Health Promotion International 34 (5): e1–17. 10.1093/heapro/day062.

Toro-Morn, M.I. (2012). Familismo. In L. S. Editor & S. M. Editor (Eds), Encyclopedia of Immigrant Health. Springer. 10.1007/978-1-4419-5659-0_277

Trauma Resource Institute. (n.d). *The community model.* https://www.traumaresourceinstitute.com/crm

Trettin, L., & Musham, C. (2000). Using focus groups to design a community health program: what roles should volunteers play?. Journal of health care for the poor and underserved, 11(4), 444–455. 10.1353/hpu.2010.0805

U.S. Department of Health and Human Services National Institutes of Health.(2021). *Clear and Simple*. https://www.nih.gov/institutes-nih/nih-office-director/office-communications-public-liaison/clear-communication/clear-simple

Verbi Software Consult. Sozialforschung GmbH. (2015). MAXQDAplus (Verson 12) [Computer software]. Berlin, Germany.

Visscher, B. B., Steunenberg, B., Heijmans, M., Hofstede, J. M., Devillé, W., van der Heide, I., & Rademakers, J. (2018). Evidence on the effectiveness of health literacy interventions in the EU: a systematic review. BMC public health, 18(1), 1414. 10.1186/s12889-018-6331-7

Volaco, A., Cavalcanti, A. M., Filho, R. P., & Précoma, D. B. (2018). Socioeconomic Status: The Missing Link Between Obesity and Diabetes Mellitus?. Current diabetes reviews, 14(4), 321–326. 10.2174/1573399813666170621123227

Wang, R., Hempton, B.M., Dugan, J.P., & Komives, S.R. (2008). Cultural Differences: Why Do Asians Avoid Extreme Responses? Survey practice, 1, 2913.

Weiss, B.D. (2007). *Health Literacy and Patient Safety: Help Patients Understand. A Manual for Clinicians* (2nd ed.). American Medical Association Foundation and American Medical Association.

Wiggins N. (2011). Critical pedagogy and popular education: towards a unity of theory and practice. Studies in the Education of Adults, 43(1): 34–49.

Wolff, K., Cavanaugh, K., Malone, R., Hawk, V., Gregory, B. P., Davis, D., Wallston, K., & Rothman, R. L. (2009). The Diabetes Literacy and Numeracy Education Toolkit (DLNET): materials to facilitate diabetes education and management in patients with low literacy and numeracy skills. The Diabetes educator, 35(2), 233–245. 10.1177/0145721709331945

World Population Review. (2024). *US Literacy by State* 2024. U.S. Literacy Rates by State 2024 (worldpopulationreview.com)

