## Supplementary material for "Creating a novel digital intervention to bridge the organizational and personal health literacy gap by teaching the way patients learn: a process and application study": Table 1

**Table 1. Recommended Steps and Time Frames to Creating a “LACASA” Intervention***

(Allow 8 to 17 months)

1. **Understand – Listen, Learn and Engage** (2 months)

- - Tune into the local culture and find local CHW partners
- - Conduct a literature search
- - Facilitate focus group discussions and interviews
- - Summarize norms, beliefs, wants and needs of population of focus

2. **Co-create Key Intervention Components** (1-3 months)

- Gather an interdisciplinary team

- Select an intervention framework

- Develop content of new intervention

- Develop new curriculum material

**3. Pre-test and Fine Tune Material** (1-3 months)

- Pre-test new material for clarity, relevance and literacy/cultural appropriateness

- Develop and test assessment/evaluation tools

**4. Pilot-test and Evaluate Intervention** (3-6 months)

- Implement new intervention in pilot groups

- Evaluate process, outcomes and population fit

**5**. **Revise Program** (1-3 months)

- Re-assess process, outcomes and sustainability

- Revise content and format of the new intervention

**6. Launch Intervention**
